# On the corona infection model with contact restriction

**DOI:** 10.1101/2020.04.08.20057588

**Authors:** Juergen Mimkes, Rainer Janssen

## Abstract

This article presents a mathematical infection model that is designed to estimate the course of coronavirus infection in Germany for several days in advance: How many people become ill or die, what is the temporal development? If the contact restriction is perfect, then the model predicts the development of the virus infection after the initial subsidence of the infection. However, since this restriction cannot always be strictly adhered to, the model is dynamically adapted to the development. This makes it possible to estimate the number of infected people, the number of new infections and deaths in Germany about a week in advance.

## The infection model for contact restriction

At the beginning of the epidemic, only a few can be infected because there are only a few people who pass on the disease. Later, with more sick people, the number of those infected grows exponentially. At some moment, there is a turning point. The new infections slowly stop because either everyone is infected or is protected from infection by contact restriction. Eventually, the total number of people infected stops growing.

With perfect contact restriction, the course of the infection can be modelled by a simple mathematical equation (“infection model”, see appendix). If contact restriction is incomplete, the model must be dynamically adapted to the course of the infection.

## The data

Since 22 January, the Johns Hopkins University (JHU) has been publishing data on worldwide corona infection. According to these data, on 5 April, in Germany, approximately 100,000 people were infected with the corona virus and more than 1500 people died. Figure 1 shows the course of infections and deaths in Germany.

**Figure 1:**
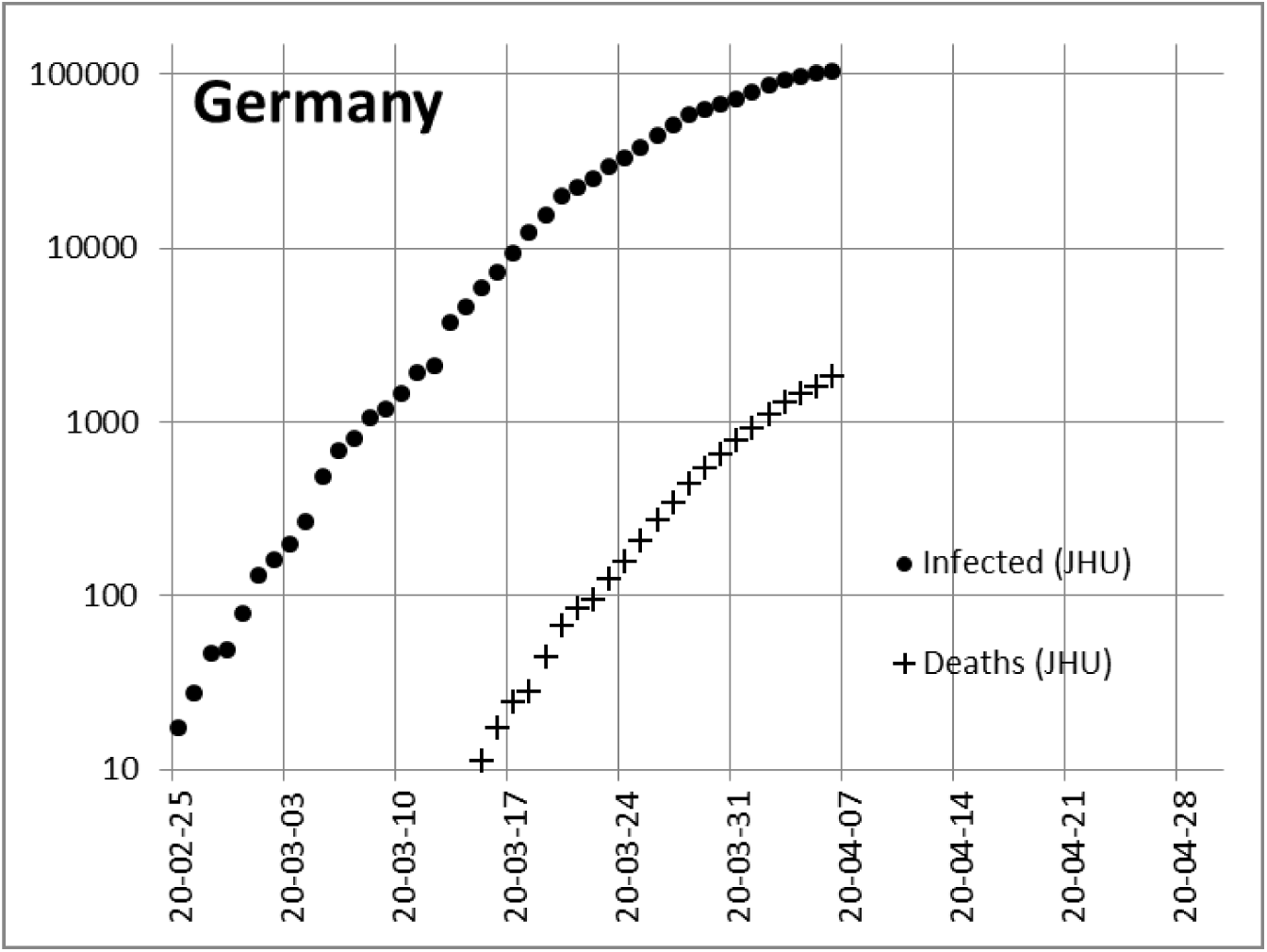
Chronological course of corona infection and deaths in Germany in logarithmic representation.

Figure 1 initially shows an exponential course of infection and deaths. In the logarithmic representation, the course is correspondingly linear. The gradient corresponds to the infection factor, if the development of the infection is unchecked. As a result of political guidelines and the behaviour of the population, the gradient has become less steep over time, i.e. the restrictions are having an effect. However, the further course of the infections and deaths is still open.

## China

To test the infection model, we have tried to model the course of infections in China after a flattening of the infection figures was observed at the end of February (Figure 2). Surprisingly, the model indicated early on that not all of Wuhan, which was already sealed off at that time, was infected with 11 million people, but only about 81,000 people. Apparently, the Chinese government has succeeded in limiting the number of infections to about 82,000 people for the time being through strict quarantine measures.

**Figure 2:**
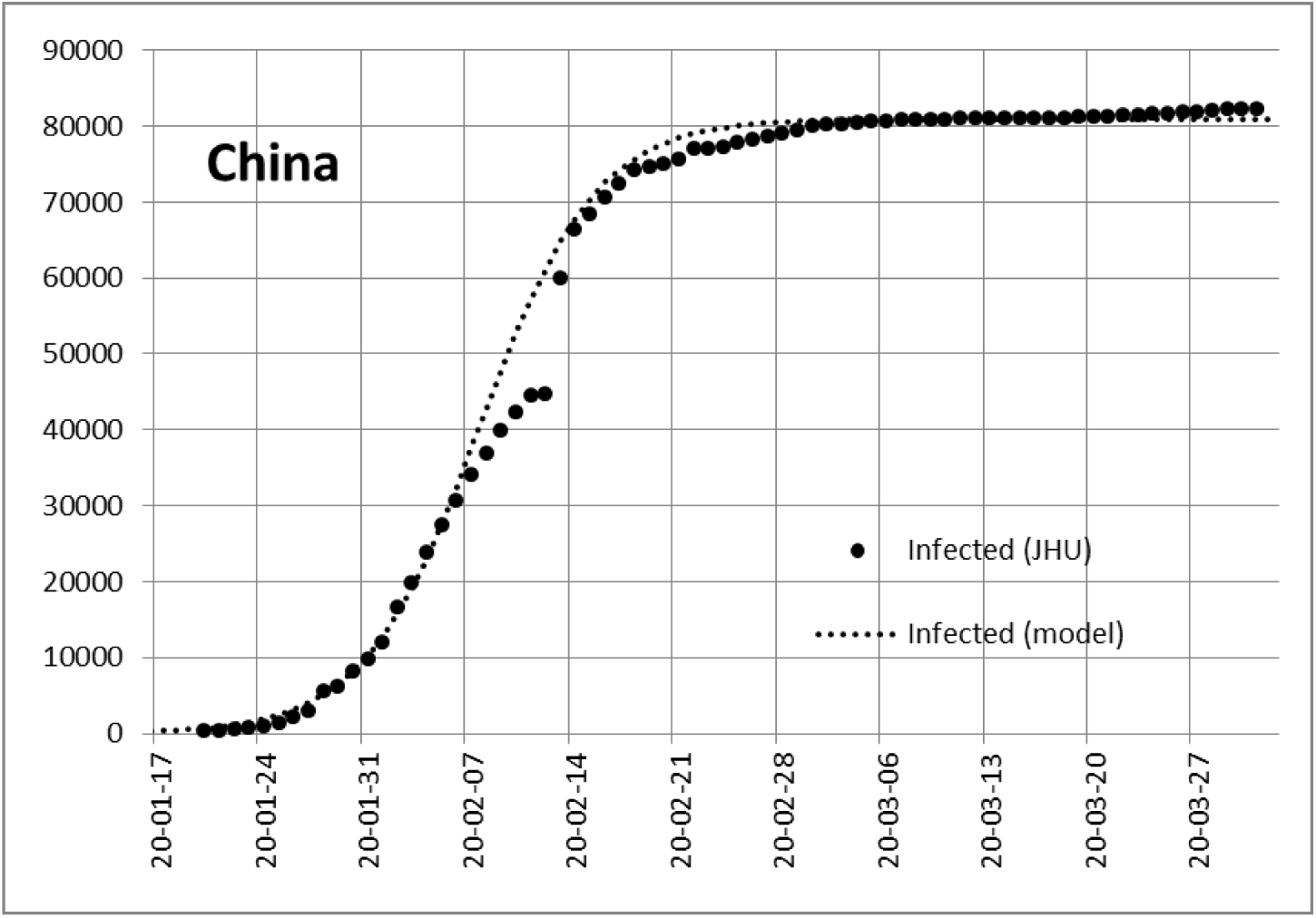
Course of corona infection in China and the adaptation of our model to the data.

## Italy

In Italy the infection figures are about a week ahead of us, so in Europe we want to examine Italy first. The number of reported infected people here had risen to about 130,000 on 5 April. Figure 3 shows the data and the development estimated according to the model, which only applies if strict isolation measures can keep the total number of infected persons at perhaps 150,000.

**Figure 3:**
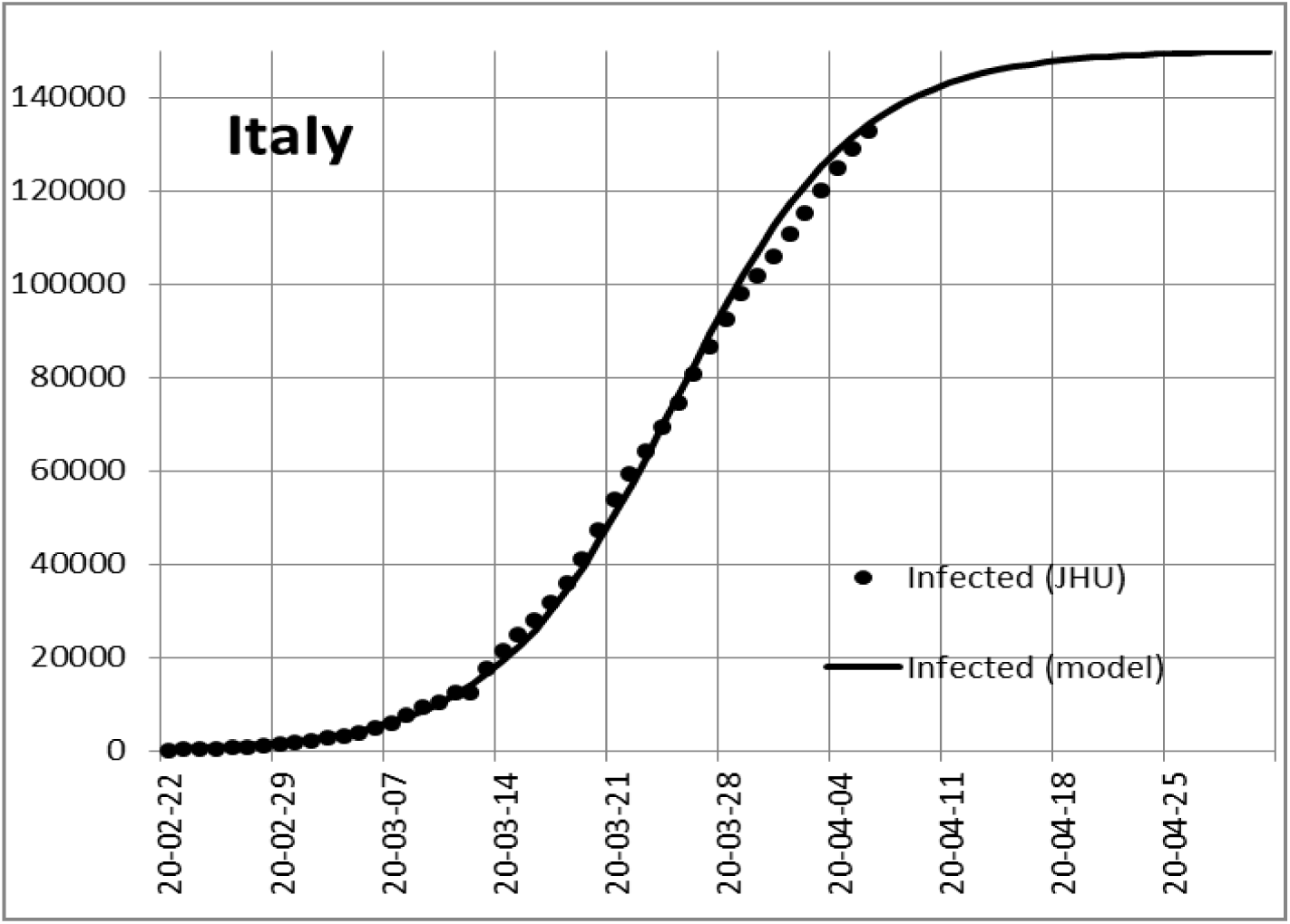
Course of corona infection in Italy and the adaptation of our model to the data.

Figure 4 shows the number of newly infected persons in Italy every day and the bell curve derived from the model as an approximation. The infection had probably reached its maximum around 25 March. It became apparent that the infection was apparently composed of three or four separate maxima, perhaps from different infection centres. However, the data of the JHU only apply to the whole of Italy and cannot take separate regions into account.

**Figure 4:**
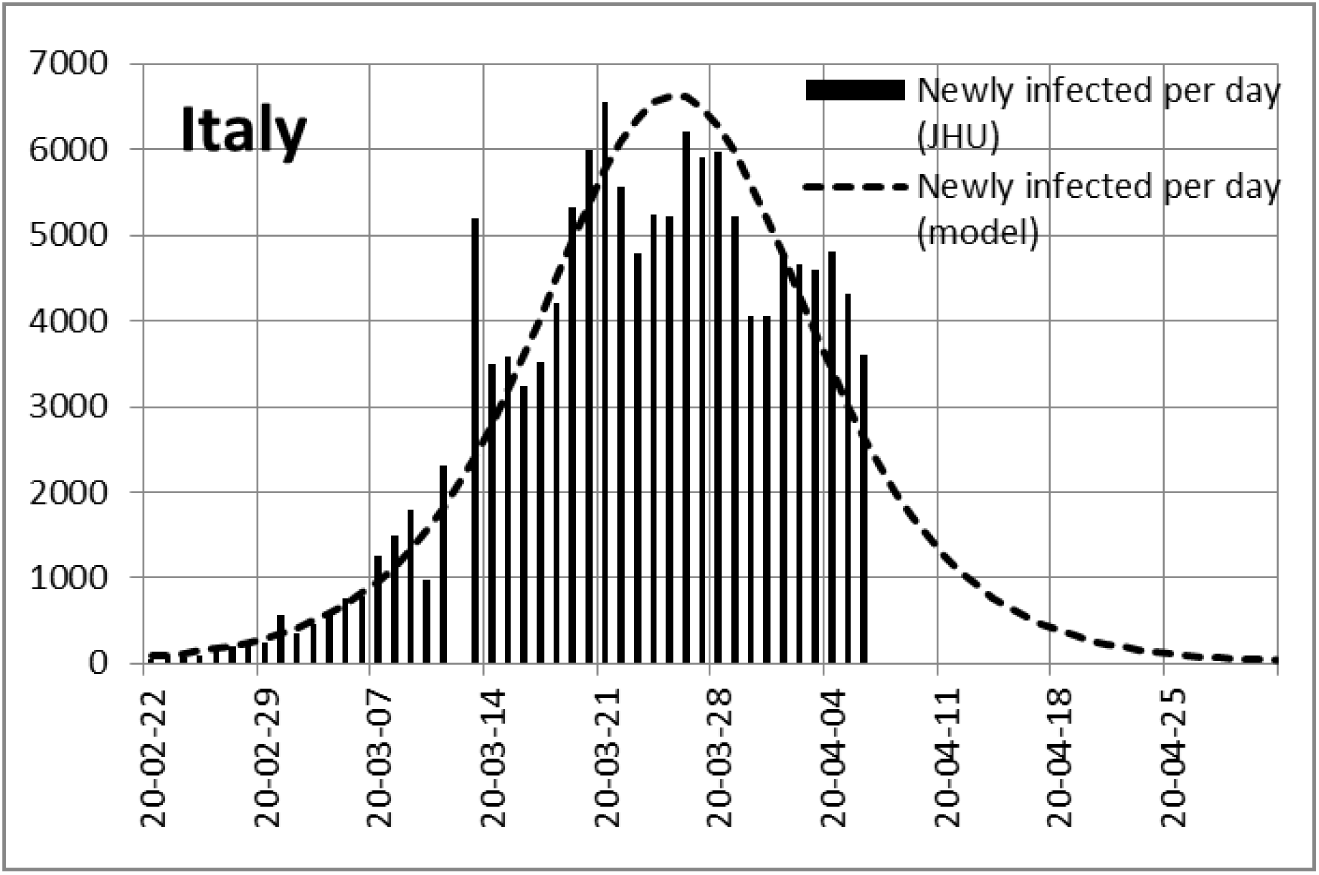
Number of daily new infections in Italy and the bell curve derived from the model as an approximation.

Figure 5 shows the number of daily deaths after JHU and the bell curve derived from the model as an approximation. The number of daily deaths had apparently reached the maximum at the end of March.

**Figure 5:**
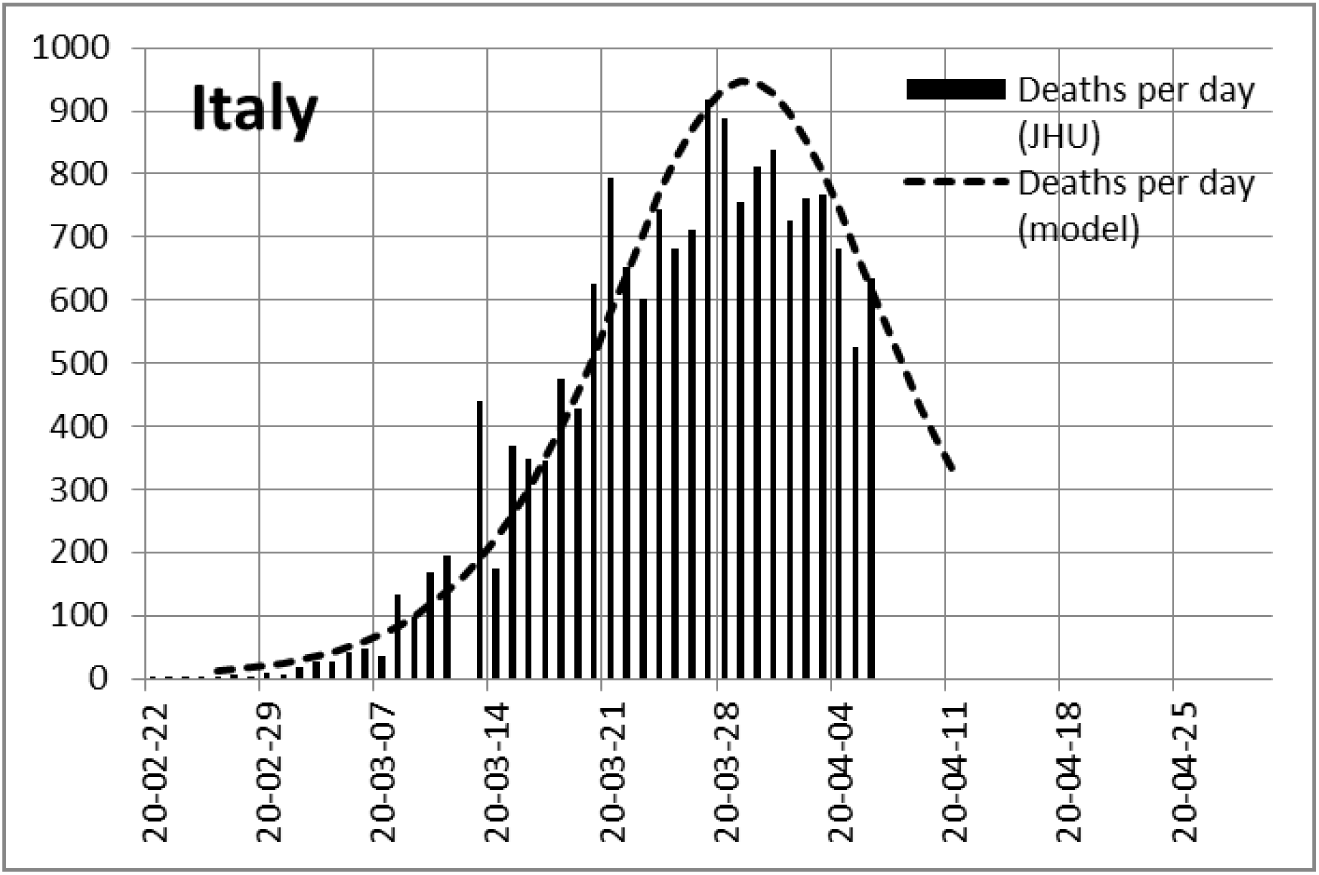
Number of daily deaths and the bell curve derived from the model as an approximation.

## Germany

Figure 6 shows the data and the possible development for a total number of patients on a linear scale (Figure 1 shows the same data on a logarithmic scale).

**Figure 6:**
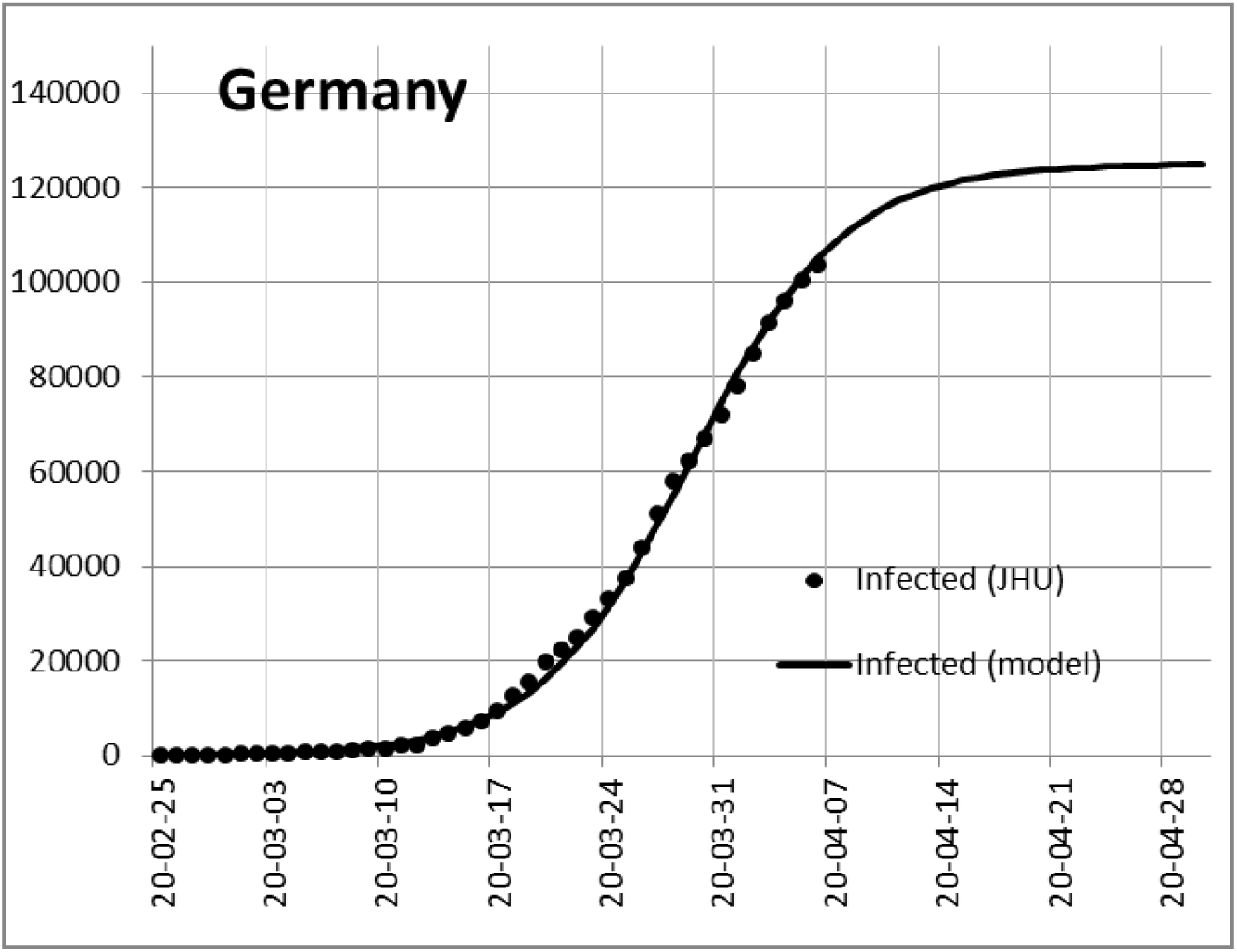
Course of the total number of all infected persons in Germany and an estimate based on the model.

The curve indicates saturation at the end of April. However, we must assume that the number may increase due to insufficient isolation of the citizens. In this way, we can achieve a forecast of perhaps a week, but we may have to adjust the course again and again to the circumstances, just as we do when driving in fog on sight.

Figure 7 indicates that the number of newly infected persons per day is decreasing. The course shows several maxima, similar to Italy. The envelope is again the model curve from the adjustment to the data in Figure 6. If the contact restriction continued to be fully effective, the minimum number of newly infected persons would probably be below 100 according to the model at the beginning of May. There is therefore a possibility that contact restrictions could be relaxed in May so that hospitals can cope with the low case numbers.

**Figure 7:**
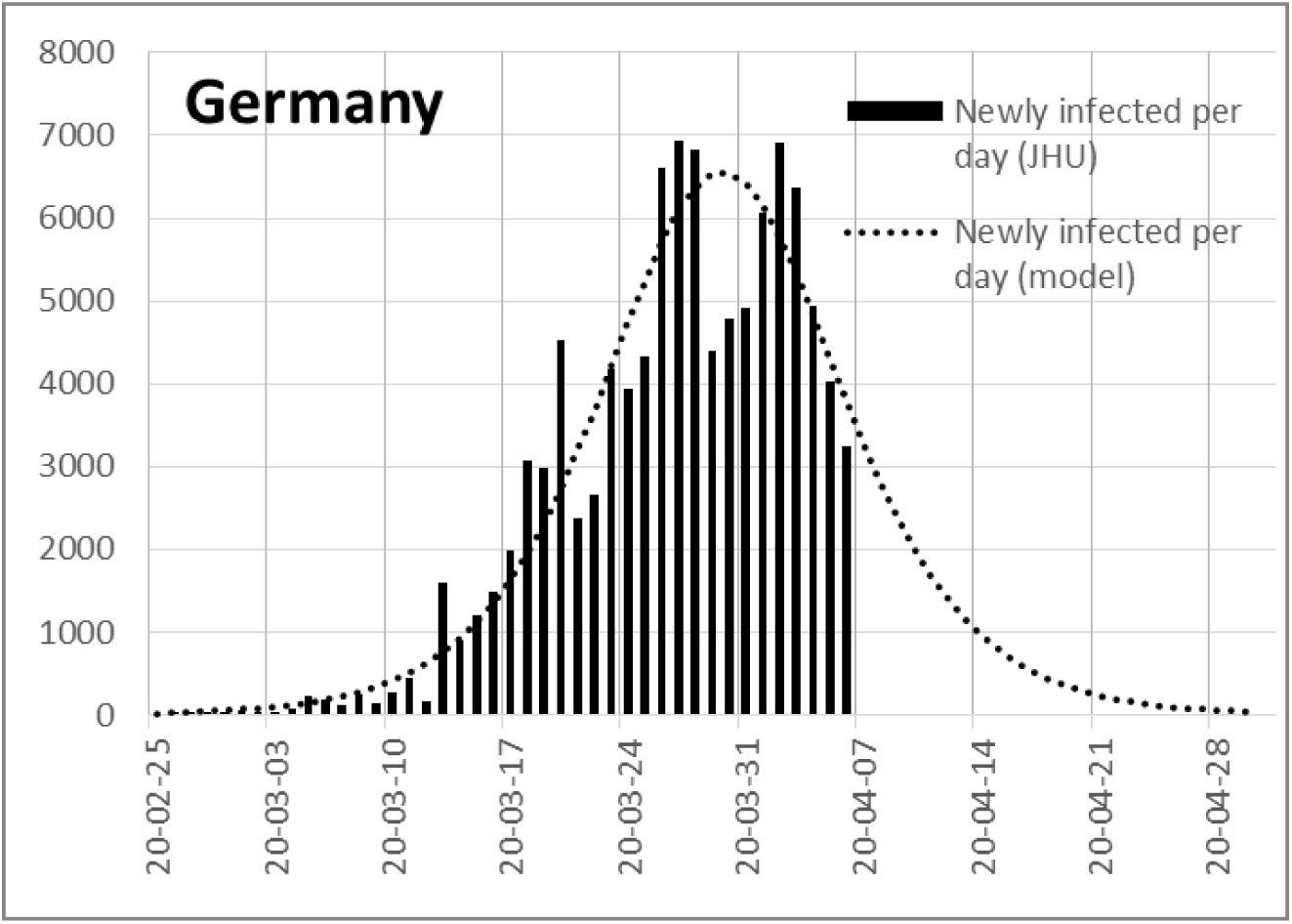
Course of the daily newly infected persons in Germany and the enveloping bell curve of the model.

Figure 8 shows the number of daily deaths and the bell curve derived from the model as an approximation.

**Figure 8:**
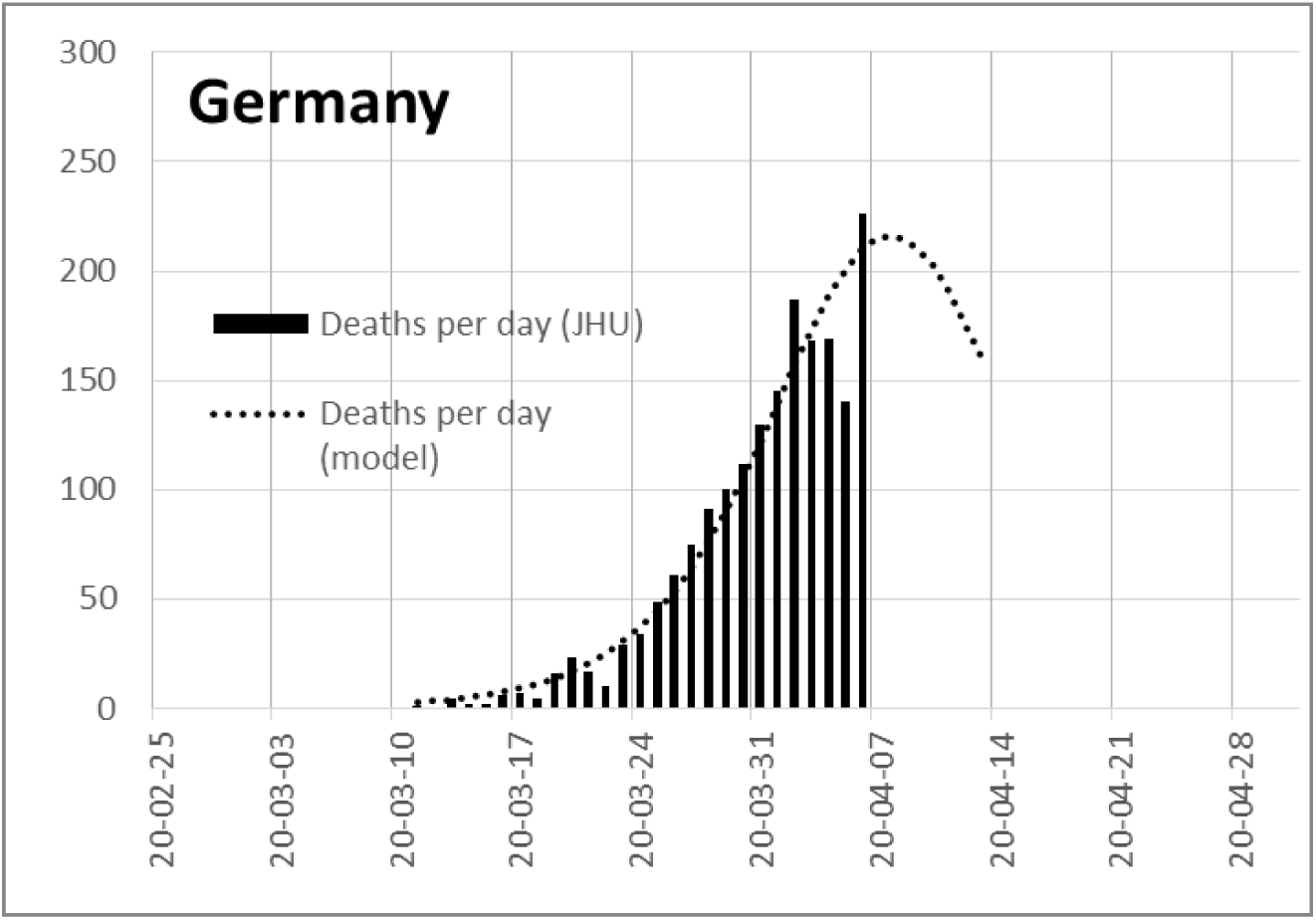
Number of daily deaths and the bell curve derived from the model as an approximation.

## Mortality

In a study (Spiegel-Online of April 2) it is assumed that the interval between real infection and date of death is 10 days in all countries. However, the number of people really infected is not known, only the number of those reported infected.

In order to examine mortality more closely, we compared the reported number of infected persons with the reported number of deaths and used this information to determine the period between infection and date of death and the mortality rate.

## Italy

For Italy, we have observed that the time between the reported number of infected persons and deaths is only about 5 days. This results in a death rate of about 14 % since the beginning of the pandemic (Figure 9).

**Figure 9:**
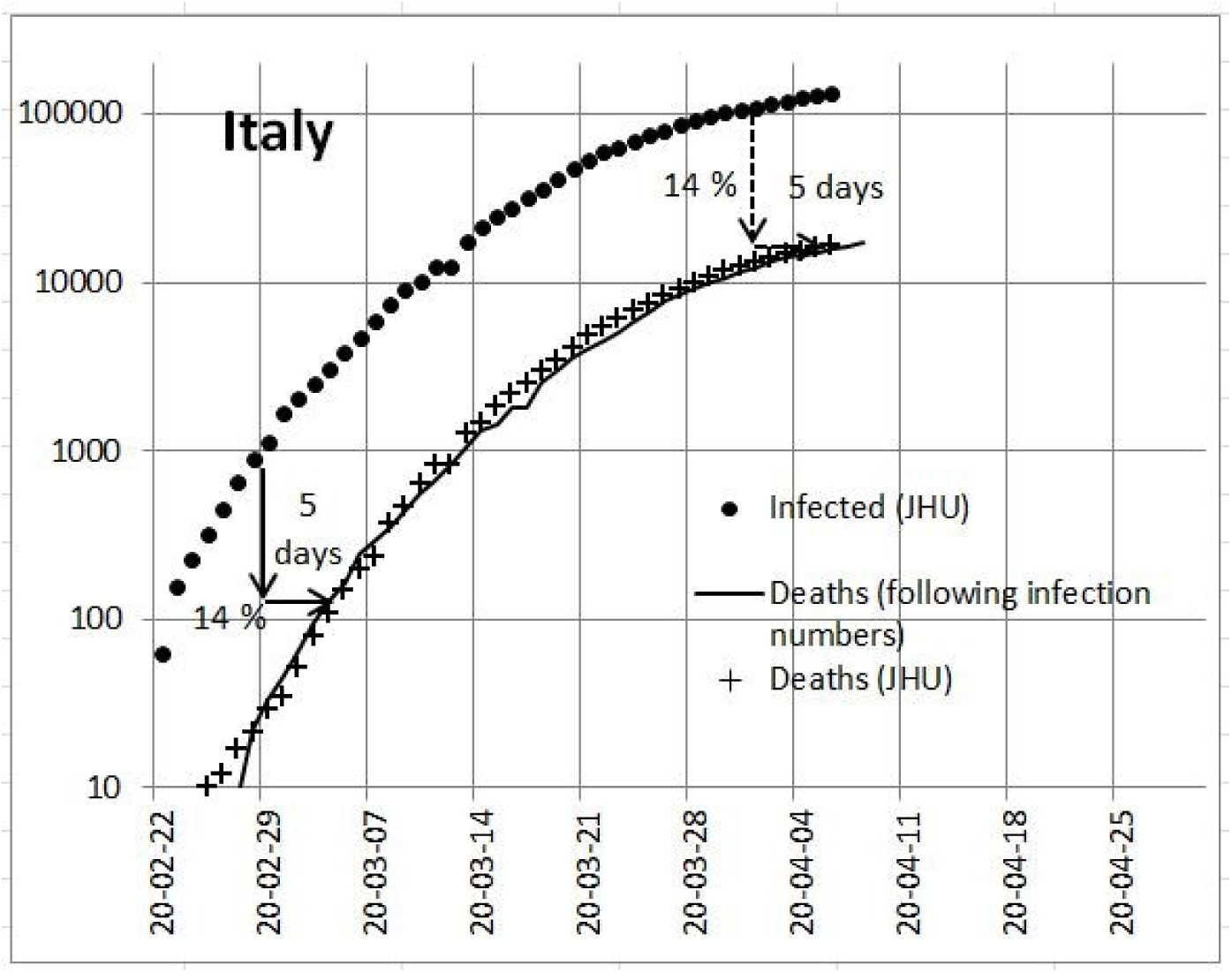
Infected and deaths in Italy

If this correlation continues to be confirmed and the methods of recording and treatment do not change, it will be possible to estimate the number of deaths at least for the next five days. In Figure 5, the 5-day intervals have already been used as an approximation for adaptation to the infection model.

## Germany

In Germany, according to Figure 10, the number of deaths seems to follow the reported number of infected persons at a distance of about 9 days. This would result in a mortality rate of about 3.5 % in Germany, which we have also used for the model in Figure 8.

**Figure 10:**
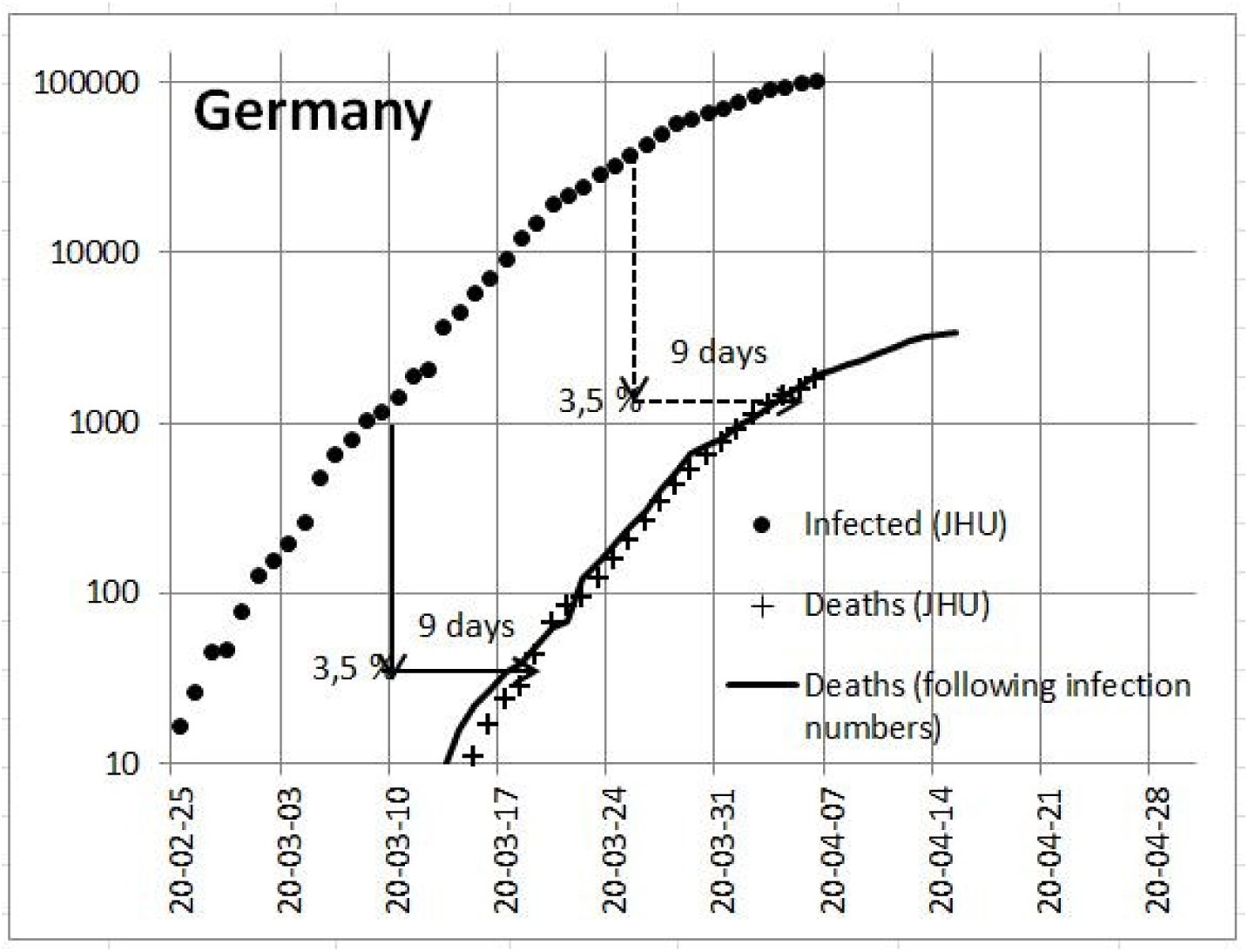
Infected and deaths in Germany

However, Figure 8 also shows that due to the time interval of nine days, the maximum number of deaths per day is still ahead of us, although the number of newly infected persons is falling. If the model is approximately correct and the methods of detection and treatment do not change, the maximum number of deaths per day could reach us in the 2nd week of April (see Figure 8).

## Sources

https://github.com/CSSEGISandData/COVID-19/tree/web-data/data

https://www.spiegel.de/wissenschaft/medizin/corona-pandemie-was-uns-die-zahl-der-toten-verraet-a-ca5dc909-716c-44ac-806f-530a10916121

## Data Availability

All data are published by Corona Virus tables by Johns Hopkins University

## Annex

### The infection model with contact restriction

The increase in infections d n(t) / d t in a country with a perfect contact ban depends on the number of people who can be infected (N) and on the number of people who fall ill n(t).

Since people who are already ill can no longer be infected, only a fraction (1 -n(t)/ N) of the population can become infected,

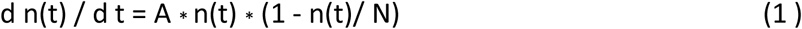

Factor (A) is the probability of infecting someone else. The solution of this equation gives

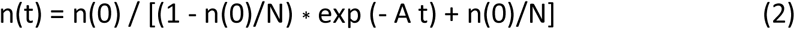

The number (t) represents the day since the beginning of the infection. With this equation the data of the corona infection for Germany were adjusted. The model only depends on three parameters: the beginning of the infection, the probability of infection and the number of all persons who can be infected. The following parameters were chosen at the beginning of April:

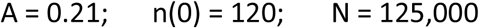

These parameters result from the representation of the infections over time.

The parameter A = 0.21 means that for 100 patients in the initial phase of the infection, 21 newly infected persons are added the next day.

The number of people who became ill at the time t = 0, n(0) = 120, results from adjusting the model to the case numbers.

The parameter N = 125,000 means that new infections stop when a total of 125,000 people have been infected.

